# *In silico* prediction of optimal multifactorial intervention in CKD

**DOI:** 10.1101/2025.02.14.25322292

**Authors:** Agnieszka Latosinska, Ioanna K Mina, Thi Minh Nghia Nguyen, Igor Golovko, Felix Keller, Gert Mayer, Peter Rossing, Jan A Staessen, Christian Delles, Joachim Beige, Griet Glorieux, Andrew L Clark, Joost P Schanstra, Antonia Vlahou, Karlheinz Peter, Ivan Rychlík, Alberto Ortiz, Archie Campbell, Harald Rupprecht, Frederik Persson, Harald Mischak, Justyna Siwy

## Abstract

**Background and Aims:** Chronic kidney disease (CKD) significantly contributes to global morbidity and mortality. Early, targeted intervention offers an ideal strategy for mitigating this burden. Peptidomic changes inform on CKD onset and progression and hold insights for treatment strategies. We investigated the molecular effects of six different therapeutic interventions *in silico* in all possible combinations on the urine peptidome, aiming to identify the most beneficial treatment for individual patients.

**Method:** This study predicted major adverse kidney events (MAKE), defined as a ≥40% decline in estimated glomerular filtration rate (eGFR) or kidney failure (median follow up time of 1.50 (95%CI 0.35, 5.0)), using the urinary peptidomic classifier CKD273 in a retrospective cohort of 935 participants. The impact of various treatments on urinary peptidomic profiles, assessed from previous studies of four different drug-based interventions (MRA, SGLT2i, GLP1-RA and ARB), one dietary intervention (olive oil) or from exercise, was applied. Treatment effects were quantified through fold changes in peptide abundance after treatment, recalibrated to align with outcomes observed in randomized controlled trials and applied to patient-specific urinary profiles, simulating intervention effects and recalculation of CKD273 scores for each individual. For combination treatments, the effects of multiple interventions were combined to model their cumulative impact.

**Results:** Simulated interventions demonstrated a significant reduction in median CKD273 scores, from 0.57 (IQR: 0.19–0.81) before to 0.039 (IQR: -0.192–0.363) after intervention (paired Wilcoxon test, P < 0.0001), when the most beneficial treatment or combination of treatments was applied individually. The combination of all available treatments was found optimal only for 17.6% of the patients and not the most frequently predicted optimal intervention. Patients with higher baseline CKD273 scores required more complex intervention combinations to achieve the greatest reduction in scores. The findings present potential individualized treatment strategies in CKD management.

**Conclusion:** This study supports the feasibility of in silico predicting effects of therapeutic interventions on CKD progression. By identifying the most beneficial treatment combinations for individual patients, this approach paves the way for precision medicine in CKD.

Graphical abstract:
Urinary peptidomics data from different types of intervention were applied in silico to predict the impact of treatment, with the extent of the impact being calibrated to fit the observed effect in large randomized trials. The personalized optimal combination of interventions was determined in 935 patients, based on the impact on CKD273. The results indicate that guiding personalized treatment based on urine peptide data is possible and may substantially improve patient management.

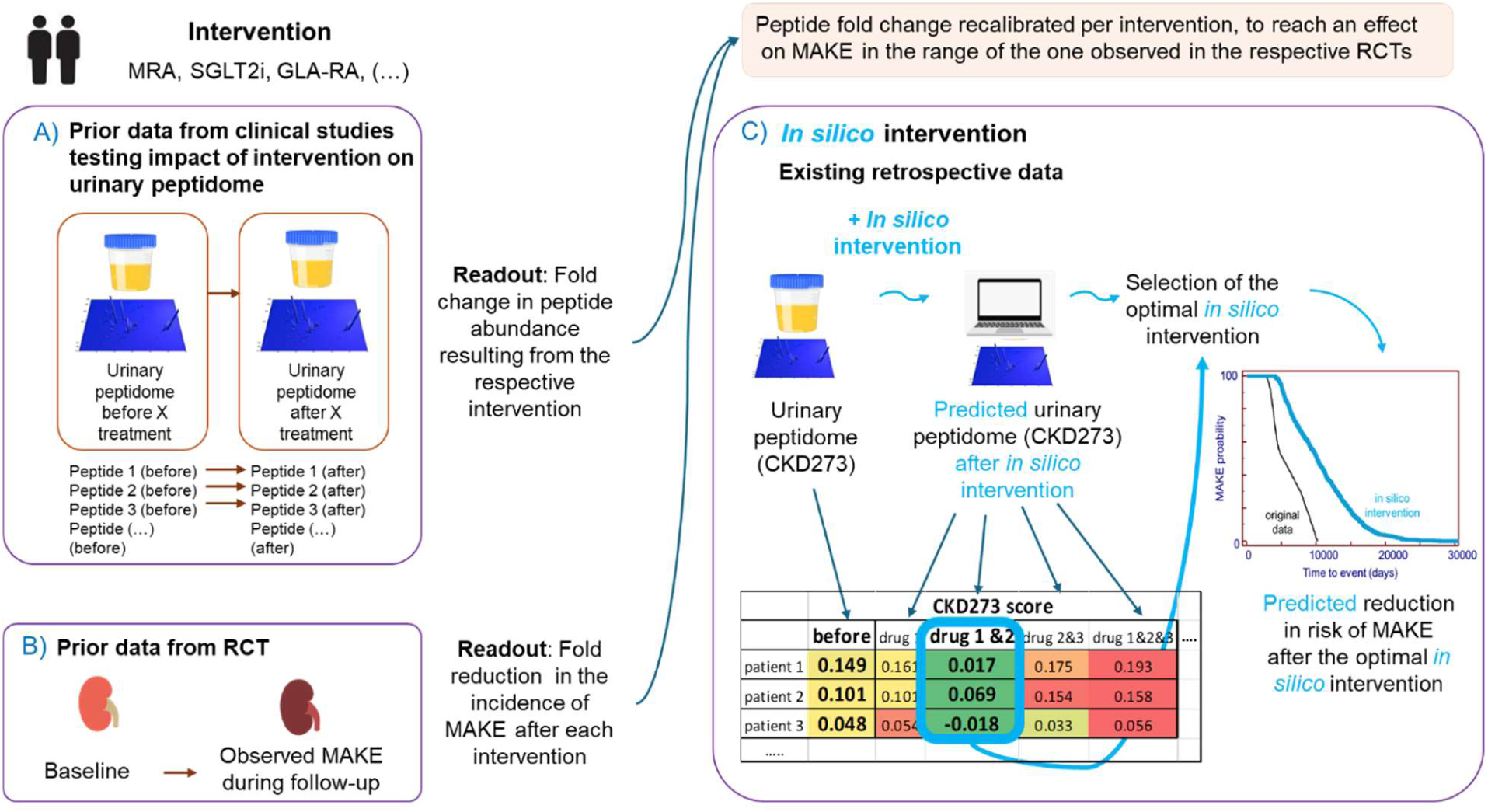

## Introduction

The development of multiple different treatment options for chronic kidney disease (CKD) has resulted in a major change of patient management. In the past, treatment was primarily focused on managing the underlying risk factors, such as hypertension and diabetes, through the use of antihypertensive and anti-diabetic therapies. Angiotensin-converting enzyme inhibitors (ACE inhibitors) and angiotensin receptor blockers (ARBs) were commonly prescribed to control blood pressure, one key driver of CKD progression. ^1^ These treatments, while beneficial in slowing disease progression in some cases, are not addressing the complex pathophysiology of kidney damage. Recently, multiple novel drug classes were introduced, all demonstrating benefit in reducing progression in CKD. Sodium-glucose co-transporter 2 inhibitors (SGLT2i), initially developed for the management of type 2 diabetes, reduce albuminuria, protect against decline in kidney function, and improve cardiovascular outcomes in patients with CKD, both with and without diabetes. ^2–4^ These effects are thought to be mediated by mechanisms such as improved glomerular hemodynamics, reduced inflammation, and enhanced tubuloglomerular feedback. ^5^ Another recent development is the use of the non-steroidal mineralocorticoid receptor antagonist (MRA), finerenone, ^6^ which targets the mineralocorticoid receptor more selectively than traditional MRAs such as spironolactone. MRAs may help counteract the fibrosis and inflammation seen in CKD progression.^7^ Glucagon-like peptide-1 (GLP-1) receptor agonists, such as semaglutide, reduce albuminuria and improve kidney outcomes, particularly in obese patients with diabetes. ^8^ Their effects are thought to be due to a combination of metabolic benefits, especially weight loss and improved insulin sensitivity. Endothelin-1 receptor antagonists are another class of drugs under investigation for CKD treatment. Endothelin-1 is a potent vasoconstrictor that contributes to kidney fibrosis and glomerular injury. Blocking the endothelin receptor, endothelin-1 antagonists, in combination with ARBs, may reduce kidney injury and progression in various forms of CKD. ^9^ Initial findings suggest that endothelin-1 antagonists could offer additional benefits for patients with specific CKD phenotypes. Additional evidence also supports non-drug interventions such as dietary and lifestyle changes in the management of CKD.^10;11^

These multiple treatment options highlight the need for specific, personalized treatment guidelines for CKD. Personalized treatment based on specific molecular features is by now routine in oncology, ^12^ with treatment targeting specific molecular changes. In contrast, due in part to the absence of a specific mutation causing CKD (with some rare exceptions like e.g. autosomal dominant polycystic kidney disease or Alport syndrome), personalized interventions are generally not implemented in nephrology. While pharmaceutical companies, in their own interest, often promote a “one-size-fits-all” approach, advocating the use of their respective drug in every patient, this strategy leads to an increased risk of adverse drug interactions, higher healthcare costs (e.g. yearly costs of Sparsentan, for the treatment of primary immunoglobulin A nephropathy, exceed 50,000 €, ^13^), and potential side effects, especially in a population already at high risk of comorbidities. Moreover, clinical evidence supporting the combined use of all these agents is generally moderate, inconclusive or non-existent. For example, studies examining the combination of GLP-1 receptor agonists with SGLT2i have failed to demonstrate an added benefit in kidney protection, ^8^ raising questions about the optimal use of combination therapies in CKD management.

We have developed a urine peptide-based classifier, CKD273, which allows for early detection of CKD and provides prognostic information regarding disease progression. ^14;15^ This classifier uses peptidomic analysis of urine to capture molecular signatures of kidney dysfunction and predict outcomes. In parallel, we have investigated how various therapeutic interventions affect the urine peptidome. ^16–22^ Using computational models, prediction of the effects of individual treatments was demonstrated as a feasible approach. ^23^ The data generated from these studies indicate substantial variability in treatment responses in individuals, underscoring the need for personalized treatment strategies. What works best for one patient may not be effective in another, even if their clinical phenotype is overall quite similar.

Here we describe an approach developed based on computational modelling and urine biomarkers to generate a predictive framework that enables clinicians to design personalized treatment regimens that maximize therapeutic efficacy while minimizing the risk of adverse effects. We investigated the molecular effects of six different therapeutic interventions in all possible combinations on the urine peptidome, with the aim of identifying the most beneficial combination of treatments for each individual patient.

## Materials and Methods

### Study participants and study design

This study includes datasets from 935 patients from previous studies on diabetes and CKD: PRIORITY, DIRECT, FLEMENGHO, CACTI, Generation Scotland, PREDICTIONS, HOMAGE, SUNmacro, and Ghent-UH. Detailed information on the designs and the methods used in these studies are available in previous publications.^15;24–30^ Inclusion criteria were availability of relevant demographic and clinical variables including age, sex, body mass index, blood pressure, estimated glomerular filtration rate (eGFR, calculated using the CKD Epidemiology Collaboration, CKD-EPI, 2009 formula) increased risk of CKD based on CKD273 scoring >0 at the time of the baseline assessment, and availability of follow-up information (eGFR or kidney failure at follow-up). The endpoint was major adverse kidney event (MAKE), defined as a decline of ≥40% in eGFR values during follow-up or kidney failure. The study was conducted according to the guidelines of the Declaration of Helsinki. All datasets were fully anonymized and from previous studies. The ethics committee of the Hannover Medical School Germany waived ethical approval under the reference number 3116-2016 for all studies involving re-use of data from anonymized urine samples.

### Peptide-based classifiers and prediction of events

The previously developed classifier CKD273 was used for prediction of CKD events and assessment of the impact of different interventions and their combinations. The scores for the classifier were predicted by a support vector machine (SVM) algorithm, integrated into the MosaDiag software,^31^ in which the levels of 273 CKD biomarkers detected in the urine of individual patients are compared to CKD-specific biomarker levels. ^14;32^ Sample classification was conducted at baseline and after *in silico* intervention. All statistical tests were performed in MedCalc version 12 - © 1993-2013 MedCalc Software.

### In-silico impact of intervention

We assessed the impact on urinary peptidomic profiles of four different drug-based interventions (MRAs, SGLT2i, GLP1 receptor agonists, and ARBs); one dietary intervention (olive oil); and one lifestyle intervention, exercise. The data on the treatment induced impact were generated in previous studies either published ^16–22^ or unpublished (exercise). Treatment effect on peptide abundance was quantified by the fold changes, given in **Supplementary Table 1**. Fold changes were calculated by dividing the average peptide abundance after treatment by the average peptide abundance before treatment (representing a result of the intervention). Missing abundance values, in general the result of the signal below detection limit, were replaced by zero. Since the follow-up after treatment was in the range of 6 weeks to 9 months, but in general not until MAKE occurred and to limit overestimation of the treatment effect, the applied fold-changes were recalibrated per treatment, ensuring that the effect on MAKE was within the range observed in the respective randomized controlled trials. ^2;6;8;33^ To simulate the effect of intervention, these re-calibrated fold changes were then applied to multiply the intensities of the respective peptides in each patient, and the predictor (CKD273) scores were re-calculated. When applying a combination of different interventions, similar principle was followed, with the effects combined (via multiplication of all relevant fold changes) **(Figure 1)**.

**Figure 1:**
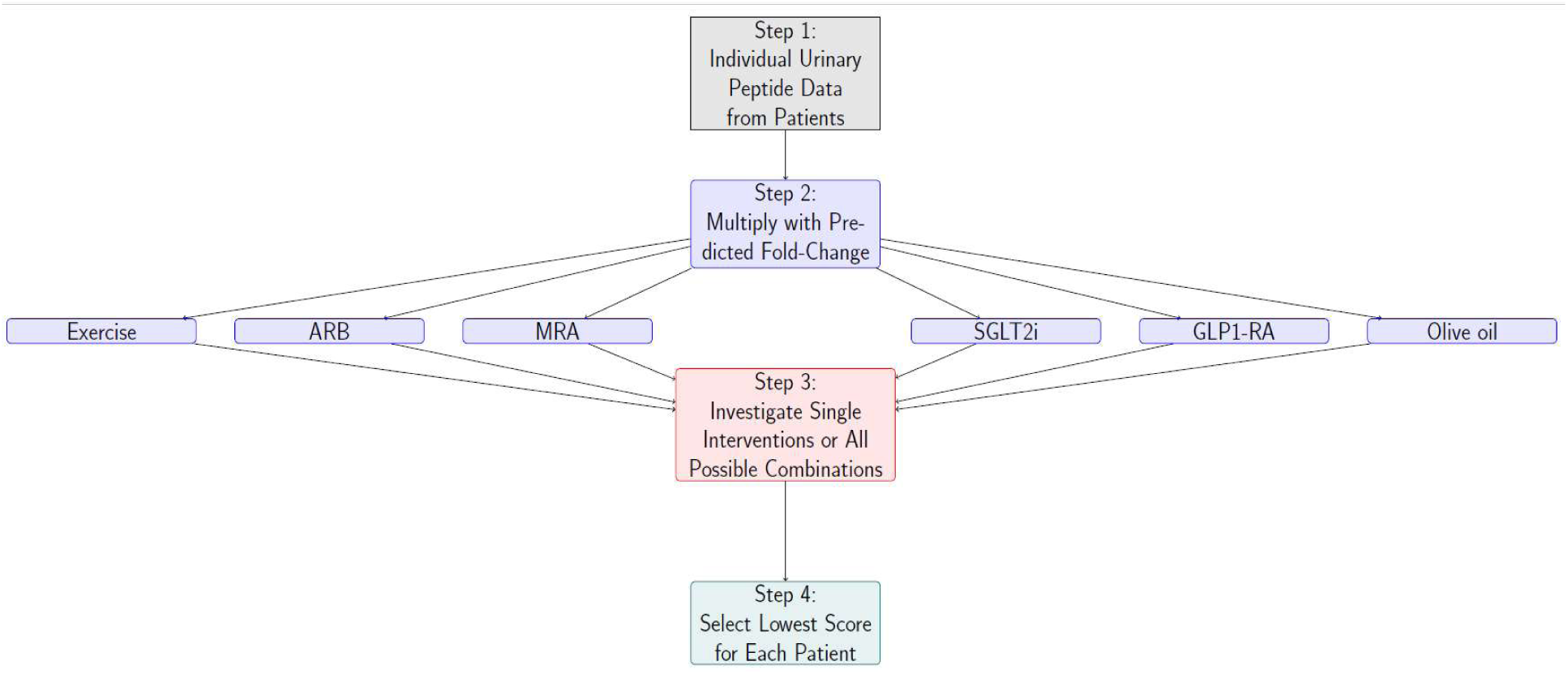
Graphical depiction of the study outline. First, urinary peptide data are acquired (Step 1), which then undergo in silico treatment using single (Step 2) or multiple (Step 3) interventions. In the last step the optimal intervention, resulting in the maximal lowering of the risk, is chosen.

### Transformation of the CKD273 biomarker score into risk of MAKE

To make CKD risk prediction more tangible and link it to a measure of risk of MAKE, the CKD273 score was transformed into the incidence rate per 100 person-years of MAKE (based on the data available from ^23^)(**Supplementary Figure 1**). For each CKD273 score, the incidence rate per 100 person-years was calculated as follows: 100 subjects with the closest CKD273 scores were selected for each target score, comprised of 50 individuals with scores above and 50 with scores below the target score value. For each subset of 100 subjects, the observed frequency of MAKE events was divided by the sum of observation times. Accordingly, the expected time to observe 50% MAKE events in a cohort with a specific CKD273 score was derived. For a detailed example, see also **Supplementary Table 2**.

The relationship between the CKD273 score and the estimated time to a 50% event rate of MAKE was then assessed using regression analysis to establish the following predictive model: **Log “time-to-50%-event-rate” = 4.1446-0.9498*CKD273 score.** The derived formula was subsequently used to transform both original CKD273 scores and scores modified after *in silico* interventions.

## Results

### Cohort characteristics

Based on the applied criteria, 935 datasets were retrieved, corresponding to 935 individuals. The subjects were predominantly male (73%), diabetic (97.8%) and obese (median BMI: 30 kg/m^2^). They had a median age of 63 years and impaired kidney function (median eGFR: 46 mL/min/1.73 m²). The median CKD273 score was 0.57 (95% CI: 0.02, 1.02). The summary clinical and demographic variables are listed in **Table 1**, and in **Supplementary Table 2** on an individual level.

**Table 1.**
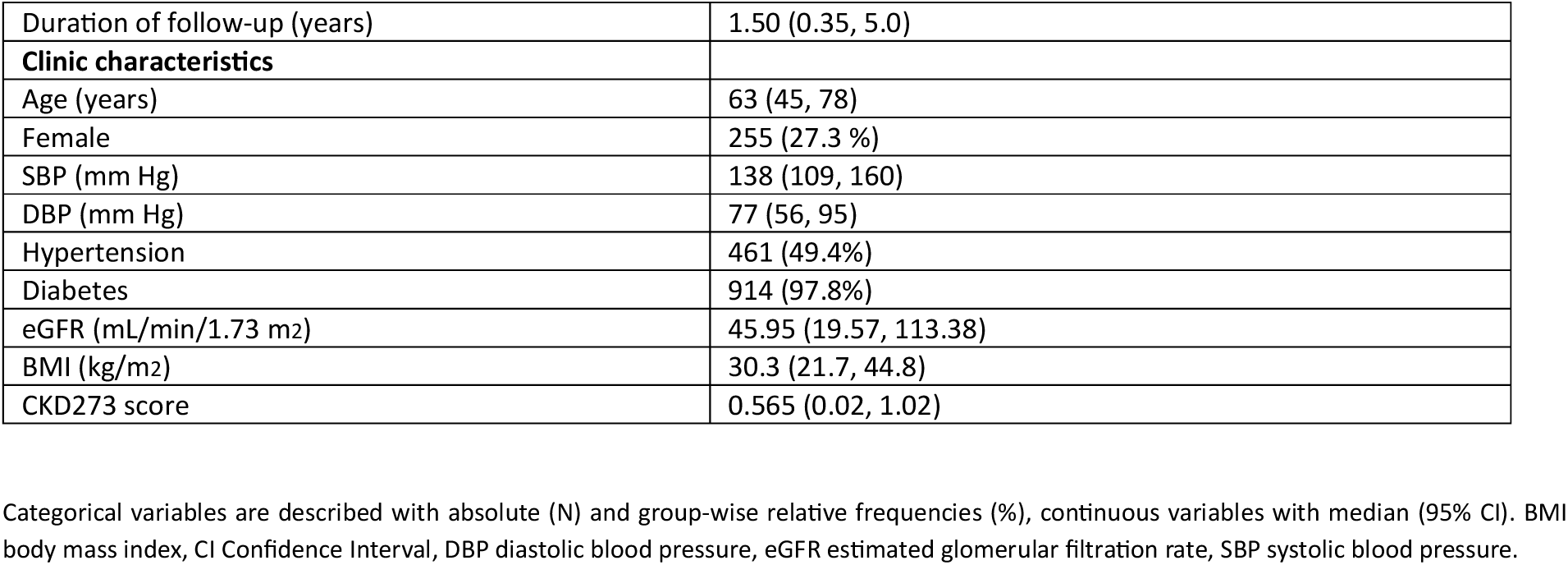
Descriptive statistics for the samples (n=935) analysed within the study.

### CKD273 scoring

The CKD273 score was assessed for patients at baseline and following *in silico* treatment. Each single intervention and all possible combinations were investigated. In total, this process resulted in the *in silico* application of 63 different therapeutic regimens.

For the *in silico* -treated subjects, the intervention yielding the lowest CKD273 score was selected as the optimal intervention for maximizing CKD risk reduction. CKD273 scores before and after the *in silico* interventions are detailed in **Supplementary Table 3**. Following the predicted most beneficial intervention, there was a reduction in the median CKD273 score from 0.57 before treatment to 0.039 after treatment (paired Wilcoxon test, P < 0.0001) (**Figure 2A**). There was a clear shift in the distribution of scores toward negative values (**Figure 2B**), indicating a potential reduction of CKD progression. For every subject, at least one intervention or combination of interventions successfully reduced the CKD273 score. The mean decrease in CKD273 score (delta) before and after treatment, based on the lowest score achieved, was 0.45.

**Figure 2:**
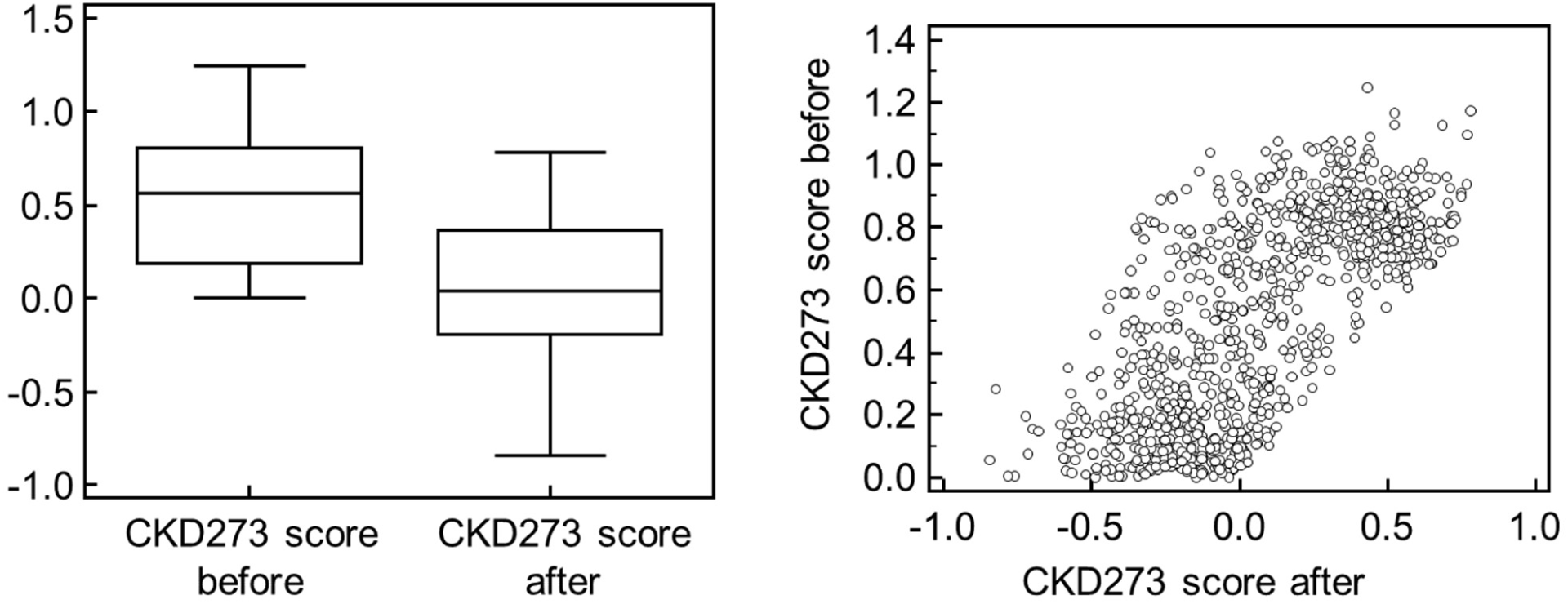
Assessment of CKD273 scores before and after in silico intervention. Box-and-whisker plots (A) and scatter plot (B) illustrating the distribution of CKD273 scores before and after the in silico intervention. The scatter plot represents CKD273 scores (before) in the horizontal axis and CKD273 scores (after) in the vertical axis. Note the different scale of the two axes. For in silico-treated subjects, the intervention that resulted in the lowest CKD273 score was plotted.

The number of patients benefiting most (based on the largest reduction in CKD273) from a specific intervention (single or combination) is shown in **Table 2**. Interestingly, the combination of all available treatments was not the most frequently predicted as optimal intervention. Instead, specific combinations, such as MRA + SGLT2i + ARB, were frequently predicted as optimal interventions compared to single or double interventions. Patients for whom *in silico* interventions predicted treatment with a higher number of drugs tended to have higher CKD273 scores before treatment (**Figure 3**). This suggests that individuals with a higher initial risk required more complex combinations of interventions to achieve the maximal reduction in CKD273 scores.

**Figure 3:**
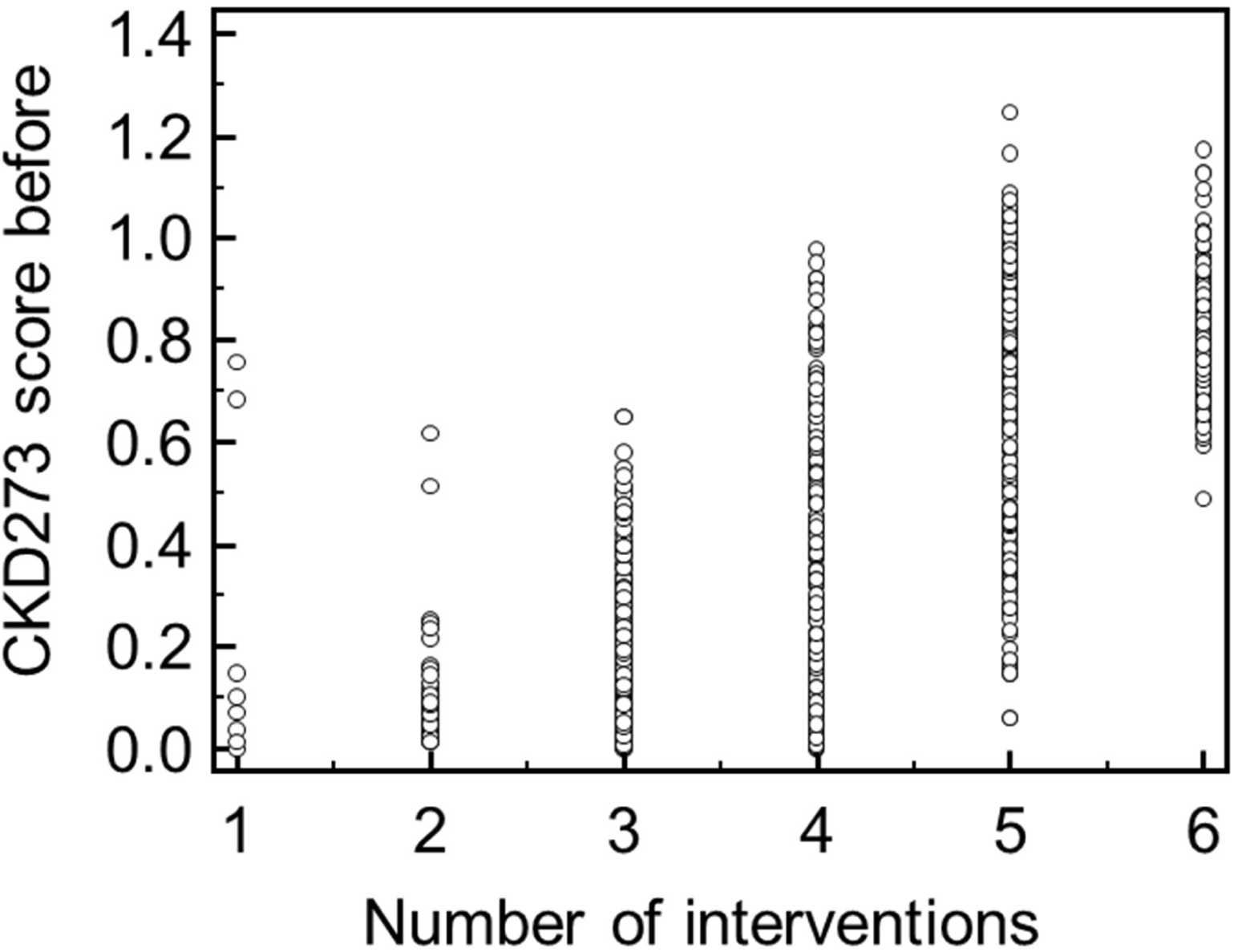
Baseline CKD273 score vs. predicted intervention count. Plot illustrating the association between the CKD273 score of patients prior to the intervention and the number of interventions predicted to be applied in combination to result in the lowest CKD273 score post-intervention.

**Table 2.**
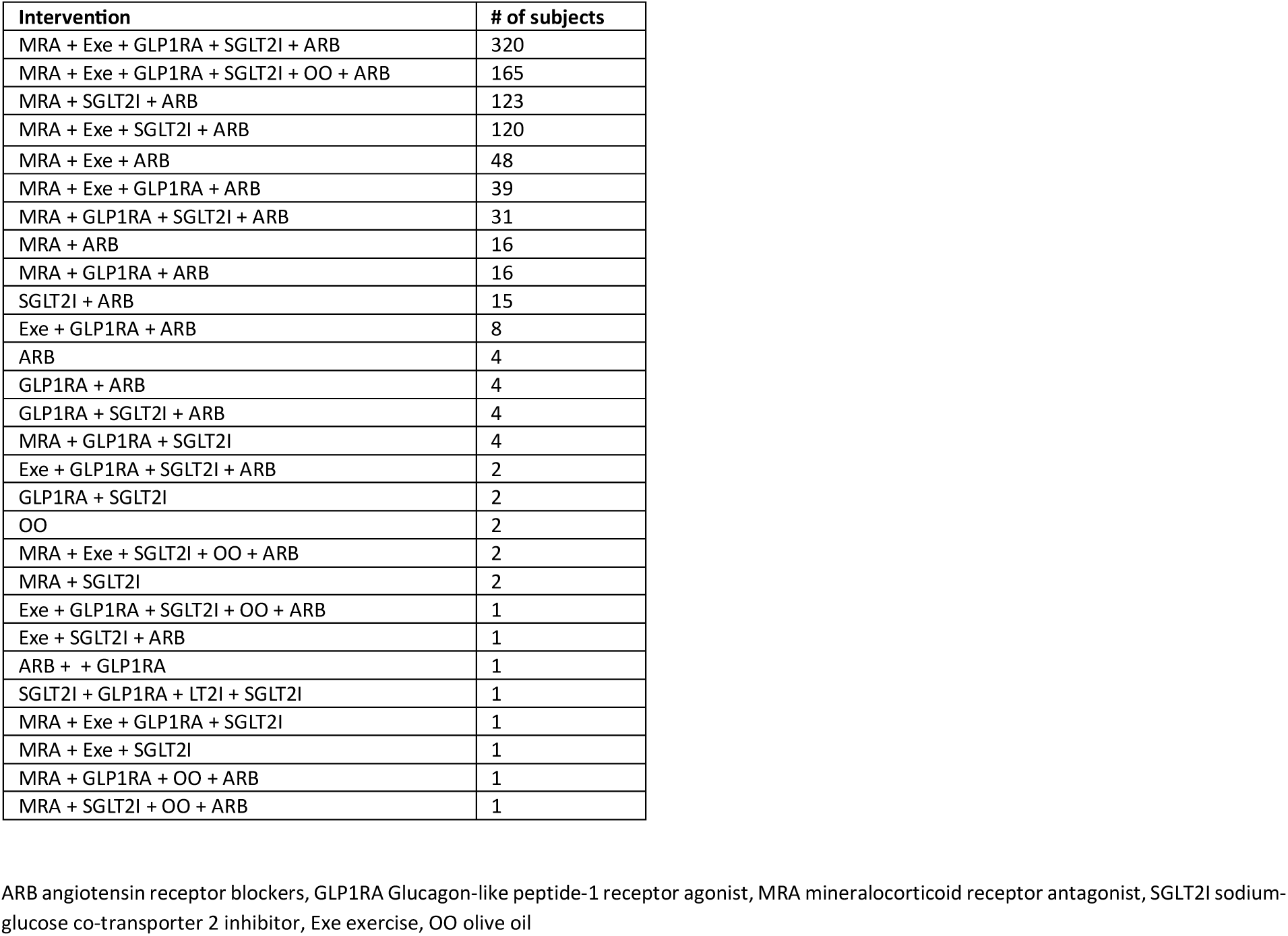
Overview of the number of subjects, predicted optimal interventions, and their combinations that resulted in the maximal reduction of risk.

| Intervention | # of subjects |
| --- | --- |
| MRA + Exe + GLP1RA + SGLT2I + ARB | 320 |
| MRA + Exe + GLP1RA + SGLT2I + OO + ARB | 165 |
| MRA + SGLT2I + ARB | 123 |
| MRA + Exe + SGLT2I + ARB | 120 |
| MRA + Exe + ARB | 48 |
| MRA + Exe + GLP1RA + ARB | 39 |
| MRA + GLP1RA + SGLT2I + ARB | 31 |
| MRA + ARB | 16 |
| MRA + GLP1RA + ARB | 16 |
| SGLT2I + ARB | 15 |
| Exe + GLP1RA + ARB | 8 |
| ARB | 4 |
| GLP1RA + ARB | 4 |
| GLP1RA + SGLT2I + ARB | 4 |
| MRA + GLP1RA + SGLT2I | 4 |
| Exe + GLP1RA + SGLT2I + ARB | 2 |
| GLP1RA + SGLT2I | 2 |
| OO | 2 |
| MRA + Exe + SGLT2I + OO + ARB | 2 |
| MRA + SGLT2I | 2 |
| Exe + GLP1RA + SGLT2I + OO + ARB | 1 |
| Exe + SGLT2I + ARB | 1 |
| ARB + + GLP1RA | 1 |
| SGLT2I + GLP1RA + LT2I + SGLT2I | 1 |
| MRA + Exe + GLP1RA + SGLT2I | 1 |
| MRA + Exe + SGLT2I | 1 |
| MRA + GLP1RA + OO + ARB | 1 |
| MRA + SGLT2I + OO + ARB | 1 |

ARB angiotensin receptor blockers, GLP1RA Glucagon-like peptide-1 receptor agonist, MRA mineralocorticoid receptor antagonist, SGLT2I sodium-glucose co-transporter 2 inhibitor, Exe exercise, OO olive oil

### Risk of MAKE based on the CKD273 scoring

**Supplementary Figure 1** displays the relationship between the estimated time to a 50% event rate of MAKE and the baseline CKD273 score for the subjects in the study. The relation was used to develop a predictive model to estimate the time to a 50% event rate of MAKE, followed by incidence rate per 100 person-years risk of MAKE, based on the CKD273 score. The respective CKD273 scoring and calculations are given in **Supplementary Table 3**. Based on this transformation, the median incidence rate per 100 person-years risk of MAKE was 4.5 and 1.43 at the baseline and after the predicted optimal intervention **(Figure 4),** with a median relative risk reduction of 61% (**Supplementary Table 3**). Briefly, if a study followed 1,000 people for one year, we would expect about 45 and 14 new cases of MAKE without and after the application of the *in silico* intervention, based on the median incidence rates.

**Figure 4:**
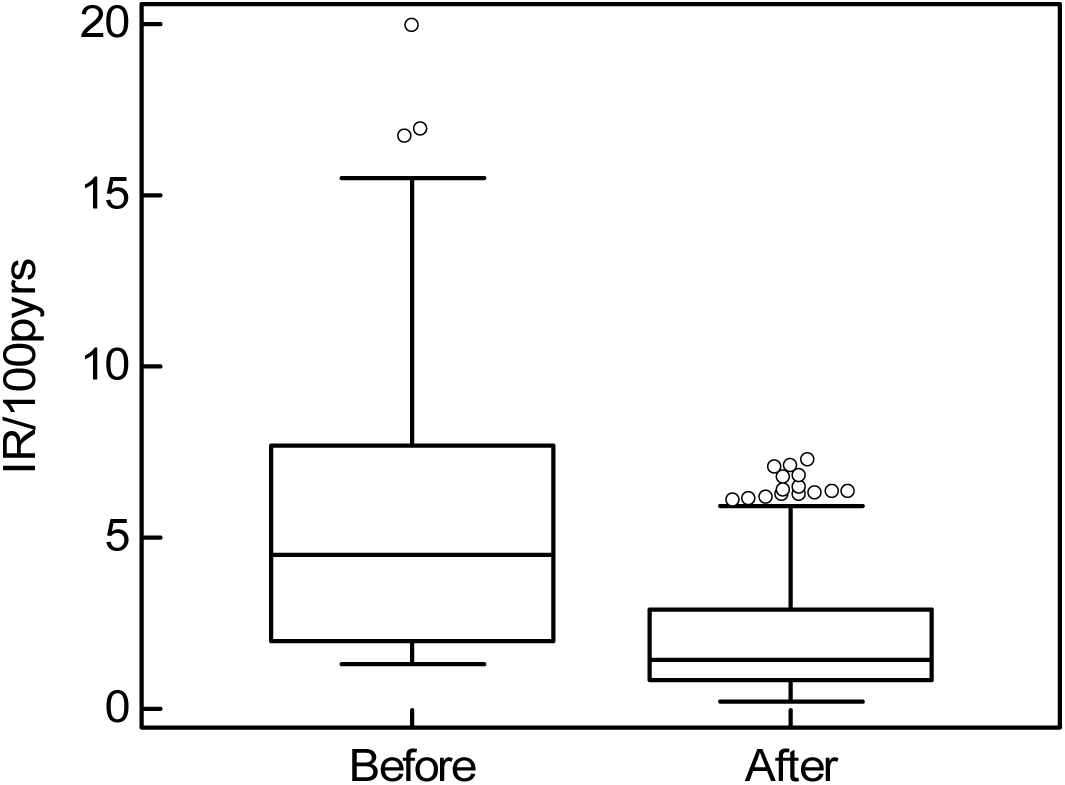
Box-and-whisker plots illustrate the incidence rate per 100 person-years before and after the in silico intervention. A significant decrease is observed following the application of the in silico intervention (paired Wilcoxon test, P < 0.0001), indicating a reduction in risk after the intervention. Abbreviations: IR incidence rate, pyers person years.

## Discussion

Prognosis of response to intervention has been identified as a key topic in patient management, with proteomics holding the promise of providing solutions. ^34^ As a first step in this direction, the PRE-score has been introduced by the group of Heerspink and colleagues to predict response to therapy, based on selected clinical variables. ^35^ The availability of a plethora of different drugs and drug classes has nurtured interest in the concept of multifactorial intervention in CKD in recent years, driven by evidence suggesting that addressing multiple pathways simultaneously can slow progression, reduce complications, and improve quality of life. The 2024 KIDGO guidelines ^36^ list 4 pharmacological interventions to treat CKD and the list is likely to grow given ongoing clinical trials and non-specific kidney protective drugs already approved for specific indications, frequently at very high cost (e.g., Nefecon for primary immunoglobulin A nephropathy is associated with yearly costs in the range of 100,000 €). However, the optimal composition and implementation of such interventions remain underexplored.

In this study we investigated an alternative approach for the management of CKD, based on molecular signatures. The results indicate that such an approach is feasible and may be used to guide intervention. While the benefit is not formally demonstrated in a prospective trial, it serves as guidance for treatment, which is currently lacking. It additionally may guide the design of future clinical trials. ^15^ CKD remains a major public health challenge, predicted to become the 5^th^ global cause of death by 2040. ^37;38^ This study addresses the urgent need for guidance on personalized interventions for CKD patients. The goal is to move away from a “trial-and-error” approach towards a precision medicine model for CKD management, where treatments are not only based on clinical features but are also guided by molecular insights specific to each patient. This is expected to significantly improve patient outcomes by tailoring interventions to the individual’s disease profile, optimizing the use of available therapies, and potentially delaying or even preventing the need for kidney replacement therapies such as dialysis. As the field of CKD treatment continues to evolve, personalized strategies based on molecular profiling, such as those derived from the urine peptidome, hold promise to become a cornerstone of clinical practice.

The theoretical background, to adjust treatment based on the individual molecular phenotype, is already in place in oncology. ^39^ Nephrology faces a similar dilemma: while multiple drugs are available, there is lack of sufficient guidance on which one to use and how to effectively combine them. ^36^ As such, it seems reasonable to start an approach in a similar direction: to obtain guidance on the optimal intervention from the patient’s molecular phenotype. Obviously, this is practically impossible based on changes in the tissue, but well applicable based on urine, which does reflect the “status” and (patho)physiology of the kidney. ^40–42^

By developing an in silico treatment profile for each patient, the effects of intended treatments and their combinations can be efficiently simulated based on retrospective proteomic substance-related outcome data. The CKD273 composite score captures the complexity of CKD by integrating multiple molecular markers, which represent different pathological mechanisms of the disease. Unlike single biomarkers, this approach provides a more comprehensive view of CKD. The ability to simulate the effects of drugs on this composite score, reflecting their influence on multiple molecular pathways, allows for predictions of therapeutic impact, based on molecular changes. The proposed approach provides information on each patient for all combinations of interventions, enabling clinicians to make tailored decisions. Even though specific interventions could be excluded based on patient characteristics, risks, or contraindications, based on all available data the optimal treatment excluding the specific intervention can still be defined.

The study has several limitations, in part also owed to its novelty:

1. The *in silico* intervention is based on previous data, many from moderate size studies. Future investigations of larger cohorts with a longer follow-up time should enable prediction of the drug effect with higher accuracy.
2. Possible confounders that may impact the drug effects are not known and were therefore not taken into account.
3. Neither albuminuria nor background treatment at the time of sampling were taken into account.
4. The approach assumes that the treatment effects are independent and additive, due to lack of data on potential interactions between interventions. All interventions were therefore considered independent from each other.
5. The study is observational, a causal relationship between peptidomic changes and clinical outcomes was not investigated and is not implied.
6. Ultimately, this is an *in silico* study, the exact benefit of the approach can only be assessed based on prospective trials, comparing clinical randomized trials with the *in silico* guided intervention.
7. The MRA tested in urinary peptidomics was spironolactone, a steroidal MRA while the approved drug to treat CKD is finerenone, a nonsteroidal MRA.

## Conclusion

In this study we demonstrated the feasibility of an approach using the individual, molecular proteomic/peptidomic signature of a patient to guide treatment in CKD. The study was based on the hypothesis that a pharmaco-omics approach, in this case based on urine peptides, should enable guiding a personalized approach towards management of CKD. An apparent major benefit of the approach is that due to the high dimensionality (over 20,000 peptides in urine are described and assessed), the theoretical benefit of not just one, but of combined therapeutic interventions can be predicted. In addition, impact of additional treatments (e.g. novel drugs) can be added to the approach, enabling expansion with moderate efforts. While the exact benefit of the approach needs to be determined in a prospective study, our data describe a potential profiling tool towards optimisation of CKD treatment. This is even more important considering that multiple drug classes are currently available for intervention, but no guidance on a choice of compounds is given. Such guidance may become even more relevant in the light of very high costs for novel drugs, already in some cases reaching 100,000 € per year.

## Data Availability

All data produced in the present work are contained in the manuscript

## Funding

This project was supported in part by the German ministry for education and science (BMBF) via UPTAKE (01EK2105A, 01EK2105B) to HM, JS and AL. Additional funding was provided by the ERA PerMed SIGNAL project (01KU2307), Accurate-CVD (ZIM-KK5560002AP3) funded by the by the BMWK (Federal Ministry for Economic Affairs and Climate Protection), European Union’s Horizon 2020 research and innovation programme under grant agreement No 848011 for the DC-ren project, and the European Union’s Horizon Europe Marie Skłodowska-Curie Actions Doctoral Networks – Industrial Doctorates Programme DisCo-1 (HORIZON – MSCA – 2021 – DN, 101072828) and PICKED (HORIZON-MSCA-2023-DN, 101168626). AO research is supported by FIS/Fondos FEDER ERA-PerMed-JTC2022 (SPAREKID AC22/00027). Comunidad de Madrid en Biomedicina P2022/BMD-7223. CIFRA_COR-CM. Instituto de Salud Carlos III (ISCIII) RICORS program to RICORS2040 (RD21/0005/0001) funded by European Union – NextGenerationEU. Mecanismo para la Recuperación y la Resiliencia (MRR) and SPACKDc PMP21/00109. FEDER funds. The project also received support from the COST Action PERMEDIK CA21165. supported by COST (European Cooperation in Science and Technology) and from the PREVENTCKD Consortium. Project ID: 101101220 Programme: EU4H. DG/Agency: HADEA. Views and opinions expressed are however those of the author(s) only and do not necessarily reflect those of the European Union or the granting authorities. Neither the European Union nor the granting authority can be held responsible for them.

## Conflicts of Interest

HM is the founder and co-owner of Mosaiques Diagnostics (Hannover, Germany). AL, TMNN, IG, IM and AL are employed by Mosaiques Diagnostics. PR has received grants from Astra Zeneca. Bayer and Novo Nordisk and honoraria (to Steno Diabetes Center Copenhagen) from Astra Zeneca. Abbott. Bayer. Boehringer Ingelheim. Eli Lilly. Novo Nordisk. Gilead. and Sanofi. AO has received grants from Sanofi and consultancy or speaker fees or travel support from Adviccene. Alexion. Astellas. Astrazeneca. Amicus. Amgen. Boehringer Ingelheim. Fresenius Medical Care. GSK. Bayer. Sanofi-Genzyme. Menarini. Mundipharma. Kyowa Kirin. Lilly. Freeline. Idorsia. Chiesi. Otsuka. Novo-Nordisk. Sysmex and Vifor Fresenius Medical Care Renal Pharma and is Director of the Catedra UAM-Astrazeneca of chronic kidney disease and electrolytes. He has stock in Telara Farma. F.P. has served as a consultant on advisory boards or as an educator for Astra Zeneca, Novo Nordisk, Sanofi, Mundipharma, MSD, Boehringer Ingelheim, Novartis, and Amgen, and has received research grants to institution from Novo Nordisk, Amgen, and Astra Zeneca. All other authors have no potential conflicts of interest.

---

**Supplementary Figure 1:**
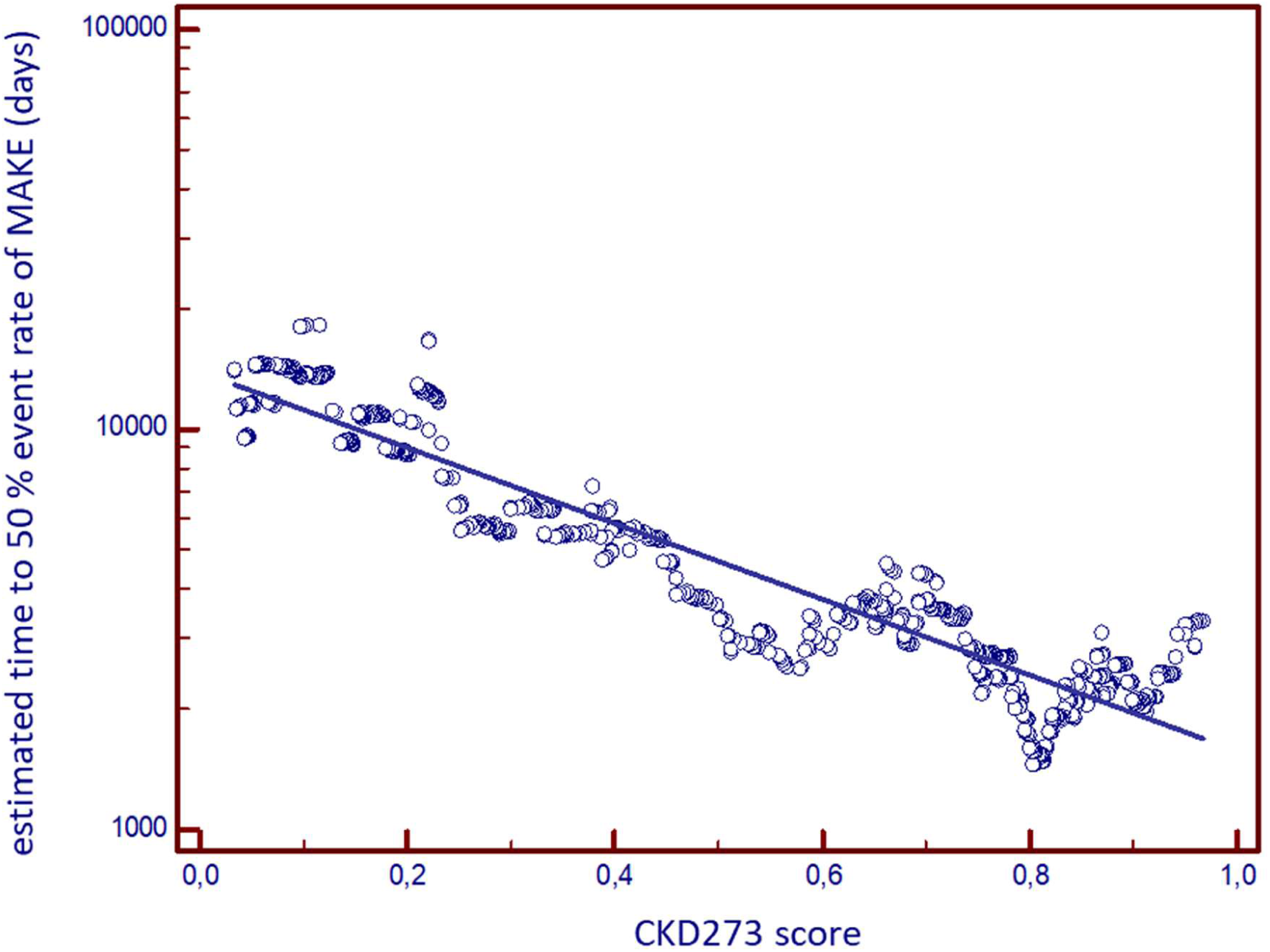
The relationship between the baseline CKD273 score and the estimated time to a 50% event rate of MAKE.

## Notes

### Author Declarations

The study used ONLY openly available human data that were originally published in Pharmaceuticals (Basel). 2023 Sep 14;16(9):1298. doi: 10.3390/ph16091298.

## Reference List

1. Vassalotti JA, Centor R, Turner BJ, Greer RC, Choi M, Sequist TD: Practical Approach to Detection and Management of Chronic Kidney Disease for the Primary Care Clinician. Am J Med 129:153–162, 2016

2. Wanner C, Inzucchi SE, Lachin JM, Fitchett D, von Eynatten M, Mattheus M, Johansen OE, Woerle HJ, Broedl UC, Zinman B: Empagliflozin and Progression of Kidney Disease in Type 2 Diabetes. New England Journal of Medicine 375:323–334, 2016

3. Neal B, Perkovic V, Mahaffey KW, de Zeeuw D, Fulcher G, Erondu N, Shaw W, Law G, Desai M, Matthews DR: Canagliflozin and Cardiovascular and Renal Events in Type 2 Diabetes. New England Journal of Medicine 377:644–657, 2017

4. Dharia A, Khan A, Sridhar VS, Cherney DZI: SGLT2 Inhibitors: The Sweet Success for Kidneys. Annu Rev Med 74:369–384, 2023

5. Bailey CJ, Day C, Bellary S: Renal Protection with SGLT2 Inhibitors: Effects in Acute and Chronic Kidney Disease. Curr Diab Rep 22:39–52, 2022

6. Bakris GL, Agarwal R, Anker SD, Pitt B, Ruilope LM, Rossing P, Kolkhof P, Nowack C, Schloemer P, Joseph A, Filippatos G: Effect of Finerenone on Chronic Kidney Disease Outcomes in Type 2 Diabetes. N Engl J Med 383:2219–2229, 2020

7. Kolkhof P, Lawatscheck R, Filippatos G, Bakris GL: Nonsteroidal Mineralocorticoid Receptor Antagonism by Finerenone-Translational Aspects and Clinical Perspectives across Multiple Organ Systems. Int J Mol Sci 23: 2022

8. Mann JFE, Rossing P, Bakris G, Belmar N, Bosch-Traberg H, Busch R, Charytan DM, Hadjadj S, Gillard P, Gorriz JL, Idorn T, Ji L, Mahaffey KW, Perkovic V, Rasmussen S, Schmieder RE, Pratley RE, Tuttle KR: Effects of semaglutide with and without concomitant SGLT2 inhibitor use in participants with type 2 diabetes and chronic kidney disease in the FLOW trial. Nat Med 30:2849–2856, 2024

9. Smeijer JD, Kohan DE, Dhaun N, Noronha IL, Liew A, Heerspink HJL: Endothelin receptor antagonists in chronic kidney disease. Nat Rev Nephrol 2024

10. Kalantar-Zadeh K, Joshi S, Schlueter R, Cooke J, Brown-Tortorici A, Donnelly M, Schulman S, Lau WL, Rhee CM, Streja E, Tantisattamo E, Ferrey AJ, Hanna R, Chen JLT, Malik S, Nguyen DV, Crowley ST, Kovesdy CP: Plant-Dominant Low-Protein Diet for Conservative Management of Chronic Kidney Disease. Nutrients 12: 2020

11. Schrauben SJ, Apple BJ, Chang AR: Modifiable Lifestyle Behaviors and CKD Progression: A Narrative Review. Kidney360 3:752–778, 2022

12. Tsimberidou AM, Fountzilas E, Nikanjam M, Kurzrock R: Review of precision cancer medicine: Evolution of the treatment paradigm. Cancer Treat Rev 86:102019, 2020

13. G-BA: https://www.g-bade/downloads/92-975-7848/2024_07_31_Modul3A_Sparsentan.pdf 2025

14. Good DM, Zürbig P, Argiles A, Bauer HW, Behrens G, Coon JJ, Dakna M, Decramer S, Delles C, Dominiczak AF, Ehrich JH, Eitner F, Fliser D, Frommberger M, Ganser A, Girolami MA, Golovko I, Gwinner W, Haubitz M, Herget-Rosenthal S, Jankowski J, Jahn H, Jerums G, Julian BA, Kellmann M, Kliem V, Kolch W, Krolewski AS, Luppi M, Massy Z, Melter M, Neususs C, Novak J, Peter K, Rossing K, Rupprecht H, Schanstra JP, Schiffer E, Stolzenburg JU, Tarnow L, Theodorescu D, Thongboonkerd V, Vanholder R, Weissinger EM, Mischak H, Schmitt-Kopplin P: Naturally occurring human urinary peptides for use in diagnosis of chronic kidney disease. Mol Cell Proteomics 9:2424–2437, 2010

15. Tofte N, Lindhardt M, Adamova K, Bakker SJL, Beige J, Beulens JWJ, Birkenfeld AL, Currie G, Delles C, Dimos I, Francova L, Frimodt-Moller M, Girman P, Goke R, Havrdova T, Heerspink HJL, Kooy A, Laverman GD, Mischak H, Navis G, Nijpels G, Noutsou M, Ortiz A, Parvanova A, Persson F, Petrie JR, Ruggenenti PL, Rutters F, Rychlik I, Siwy J, Spasovski G, Speeckaert M, Trillini M, Zurbig P, von der LH, Rossing P: Early detection of diabetic kidney disease by urinary proteomics and subsequent intervention with spironolactone to delay progression (PRIORITY): a prospective observational study and embedded randomised placebo-controlled trial. Lancet Diabetes Endocrinol 8:301–312, 2020

16. Curovic RV, Eickhoff MK, Ronkko T, Frimodt-Moller M, Hansen TW, Mischak H, Rossing P, Ahluwalia TS, Persson F: Dapagliflozin Improves the Urinary Proteomic Kidney-Risk Classifier CKD273 in Type 2 Diabetes with Albuminuria - a Randomized Clinical Trial. Diabetes Care 2022

17. Lohia S, Siwy J, Mavrogeorgis E, Eder S, Thoni S, Mayer G, Mischak H, Vlahou A, Jankowski V: Exploratory Study Analyzing the Urinary Peptidome of T2DM Patients Suggests Changes in ECM but Also Inflammatory and Metabolic Pathways Following GLP-1R Agonist Treatment. Int J Mol Sci 24: 2023

18. Jaimes Campos MA, Mavrogeorgis E, Latosinska A, Eder S, Buchwinkler L, Mischak H, Siwy J, Rossing P, Mayer G, Jankowski J: Urinary peptide analysis to predict the response to blood pressure medication. Nephrol Dial Transplant 39:873–883, 2024

19. Silva S, Bronze MR, Figueira ME, Siwy J, Mischak H, Combet E, Mullen W: Impact of a 6-wk olive oil supplementation in healthy adults on urinary proteomic biomarkers of coronary artery disease, chronic kidney disease, and diabetes (types 1 and 2): a randomized, parallel, controlled, double-blind study. Am J Clin Nutr 101:44–54, 2015

20. Andersen S, Mischak H, Zürbig P, Parving HH, Rossing P: Urinary proteome analysis enables assessment of renoprotective treatment in type 2 diabetic patients with microalbuminuria. BMC Nephrol 11:29, 2010

21. Siwy J, Klein T, Rosler M, von Eynatten M: Urinary Proteomics as a Tool to Identify Kidney Responders to Dipeptidyl Peptidase-4 Inhibition: A Hypothesis-Generating Analysis from the MARLINA-T2D Trial. Proteomics Clinical Applications 13:e1800144, 2019

22. Yu YL, Siwy J, An DW, Gonzalez A, Hansen T, Latosinska A, Pellicori P, Ravassa S, Mariottoni B, Verdonschot JA, Ahmed F, Petutschnigg J, Rossignol P, Heymans S, Cuthbert JJ, Girerd N, Clark AL, Verhamme P, Nawrot TS, Janssens S, Cleland JG, Zannad F, Diez J, Mischak H, Ferreira JP, Staessen JA: Urinary proteomic signature of mineralocorticoid receptor antagonism by spironolactone: evidence from the HOMAGE trial. Heart 110:1180–1187, 2024

23. Jaimes Campos MA, Andujar I, Keller F, Mayer G, Rossing P, Staessen JA, Delles C, Beige J, Glorieux G, Clark AL, Mullen W, Schanstra JP, Vlahou A, Rossing K, Peter K, Ortiz A, Campbell A, Persson F, Latosinska A, Mischak H, Siwy J, Jankowski J: Prognosis and Personalized In Silico Prediction of Treatment Efficacy in Cardiovascular and Chronic Kidney Disease: A Proof-of-Concept Study. Pharmaceuticals (Basel) 16:1298, 2023

24. Lindhardt M, Persson F, Zuerbig P, Stalmach A, Mischak H, de Zeeuw D, Heerspink HL, Klein R, Orchard T, Porta M, Fuller J, Bilous R, Chaturvedi N, Parving HH, Rossing P: Urinary proteomics predict onset of microalbuminuria in normoalbuminuric type 2 diabetic patients, a sub-study of the DIRECT-Protect 2 study. Nephrology Dialysis Transplantation 32:1866–1873, 2017

25. Zhang ZY, Thijs L, Petit T, Gu YM, Jacobs L, Yang WY, Liu YP, Koeck T, Zurbig P, Jin Y, Verhamme P, Voigt JU, Kuznetsova T, Mischak H, Staessen JA: Urinary Proteome and Systolic Blood Pressure as Predictors of 5-Year Cardiovascular and Cardiac Outcomes in a General Population. Hypertension 66:52–60, 2015

26. Snell-Bergeon JK, Maahs DM, Ogden LG, Kinney GL, Hokanson JE, Schiffer E, Rewers M, Mischak H: Evaluation of urinary biomarkers for coronary artery disease, diabetes, and diabetic kidney disease. Diabetes Technol Ther 11:1–9, 2009

27. Smith BH, Campbell A, Linksted P, Fitzpatrick B, Jackson C, Kerr SM, Deary IJ, Macintyre DJ, Campbell H, McGilchrist M, Hocking LJ, Wisely L, Ford I, Lindsay RS, Morton R, Palmer CN, Dominiczak AF, Porteous DJ, Morris AD: Cohort Profile: Generation Scotland: Scottish Family Health Study (GS:SFHS). The study, its participants and their potential for genetic research on health and illness. Int J Epidemiol 42:689–700, 2013

28. Alkhalaf A, Zürbig P, Bakker SJ, Bilo HJ, Cerna M, Fischer C, Fuchs S, Janssen B, Medek K, Mischak H, Roob JM, Rossing K, Rossing P, Rychlik I, Sourij H, Tiran B, Winklhofer-Roob BM, Navis GJ: Multicentric validation of proteomic biomarkers in urine specific for diabetic nephropathy. PLoS One 5:e13421, 2010

29. Jacobs L, Thijs L, Jin Y, Zannad F, Mebazaa A, Rouet P, Pinet F, Bauters C, Pieske B, Tomaschitz A, Mamas M, Diez J, McDonald K, Cleland JG, Brunner-La Rocca HP, Heymans S, Latini R, Masson S, Sever P, Delles C, Pocock S, Collier T, Kuznetsova T, Staessen JA: Heart ‘omics’ in AGEing (HOMAGE): design, research objectives and characteristics of the common database. J Biomed Res 28:349–359, 2014

30. Verbeke F, Siwy J, Van BW, Mischak H, Pletinck A, Schepers E, Neirynck N, Magalhaes P, Pejchinovski M, Pontillo C, Lichtinghagen R, Brand K, Vlahou A, De BD, Glorieux G: The urinary proteomics classifier chronic kidney disease 273 predicts cardiovascular outcome in patients with chronic kidney disease. Nephrol Dial Transplant 36:811–818, 2021

31. Mischak H, Vlahou A, Ioannidis JP: Technical aspects and inter-laboratory variability in native peptide profiling: The CE-MS experience. Clin Biochem 46:432–443, 2013

32. Schanstra JP, Zurbig P, Alkhalaf A, Argiles A, Bakker SJ, Beige J, Bilo HJ, Chatzikyrkou C, Dakna M, Dawson J, Delles C, Haller H, Haubitz M, Husi H, Jankowski J, Jerums G, Kleefstra N, Kuznetsova T, Maahs DM, Menne J, Mullen W, Ortiz A, Persson F, Rossing P, Ruggenenti P, Rychlik I, Serra AL, Siwy J, Snell-Bergeon J, Spasovski G, Staessen JA, Vlahou A, Mischak H, Vanholder R: Diagnosis and prediction of CKD progression by assessment of urinary peptides. J Am Soc Nephrol 26:1999–2010, 2015

33. Lewis EJ, Hunsicker LG, Clarke WR, Berl T, Pohl MA, Lewis JB, Ritz E, Atkins RC, Rohde R, Raz I: Renoprotective effect of the angiotensin-receptor antagonist irbesartan in patients with nephropathy due to type 2 diabetes. N Engl J Med 345:851–860, 2001

34. Pena MJ, Mischak H, Heerspink HJ: Proteomics for prediction of disease progression and response to therapy in diabetic kidney disease. Diabetologia Epub ahead of print: 2016

35. Schievink B, de ZD, Smink PA, Andress D, Brennan JJ, Coll B, Correa-Rotter R, Hou FF, Kohan D, Kitzman DW, Makino H, Parving HH, Perkovic V, Remuzzi G, Tobe S, Toto R, Hoekman J, Lambers Heerspink HJ: Prediction of the effect of atrasentan on renal and heart failure outcomes based on short-term changes in multiple risk markers. Eur J Prev Cardiol 23:758–768, 2016

36. KDIGO 2024 Clinical Practice Guideline for the Evaluation and Management of Chronic Kidney Disease. Kidney Int 105:S117–S314, 2024

37. Foreman KJ, Marquez N, Dolgert A, Fukutaki K, Fullman N, McGaughey M, Pletcher MA, Smith AE, Tang K, Yuan CW, Brown JC, Friedman J, He J, Heuton KR, Holmberg M, Patel DJ, Reidy P, Carter A, Cercy K, Chapin A, Douwes-Schultz D, Frank T, Goettsch F, Liu PY, Nandakumar V, Reitsma MB, Reuter V, Sadat N, Sorensen RJD, Srinivasan V, Updike RL, York H, Lopez AD, Lozano R, Lim SS, Mokdad AH, Vollset SE, Murray CJL: Forecasting life expectancy, years of life lost, and all-cause and cause-specific mortality for 250 causes of death: reference and alternative scenarios for 2016-40 for 195 countries and territories. Lancet 392:2052–2090, 2018

38. Burden of disease scenarios for 204 countries and territories, 2022-2050: a forecasting analysis for the Global Burden of Disease Study 2021. Lancet 403:2204–2256, 2024

39. Hoeben A, Joosten EAJ, van den Beuken-van Everdingen MHJ: Personalized Medicine: Recent Progress in Cancer Therapy. Cancers (Basel) 13: 2021

40. Mavrogeorgis E, He T, Mischak H, Latosinska A, Vlahou A, Schanstra JP, Catanese L, Amann K, Huber TB, Beige J, Rupprecht HD, Siwy J: Urinary peptidomic liquid biopsy for non-invasive differential diagnosis of chronic kidney disease. Nephrol Dial Transplant 39:453–462, 2024

41. Catanese L, Rupprecht H, Huber TB, Lindenmeyer MT, Hengel FE, Amann K, Wendt R, Siwy J, Mischak H, Beige J: Non-Invasive Biomarkers for Diagnosis, Risk Prediction, and Therapy Guidance of Glomerular Kidney Diseases: A Comprehensive Review. Int J Mol Sci 25: 2024

42. Catanese L, Siwy J, Mavrogeorgis E, Amann K, Mischak H, Beige J, Rupprecht H: A Novel Urinary Proteomics Classifier for Non-Invasive Evaluation of Interstitial Fibrosis and Tubular Atrophy in Chronic Kidney Disease. Proteomes 9:32, 2021

